# Greek High Phenolic Early Harvest Extra Virgin Olive Oil Reduces the Over-Excitation of Information Flow based on Dominant Coupling Model in patients with Mild Cognitive Impairment: An EEG Resting-State Validation Approach

**DOI:** 10.1101/2021.01.16.21249869

**Authors:** Stavros I. Dimitriadis, Christos Lyssoudis, Anthoula C. Tsolaki, Eftychia Lazarou, Mahi Kozori, Magda Tsolaki

## Abstract

**Objective:** The balance of cross-frequency coupling (CFC) over within-frequency coupling (WFC) can build a nonlinearity index (NI) that encapsulates the over-excitation of information flow between brain areas and across experimental time. The present study investigated for the very first time how the Greek High Phenolic Early Harvest Extra Virgin Olive Oil (HP-EH-EVOO) versus Moderate Phenolic (MP-EVOO) and Mediterranean Diet (MeDi) intervention in people with Mild Cognitive Impairment (MCI) could affect their spontaneous EEG dynamic connectivity.

**Methods:** Fourty three subjects (14 in MeDi, 16 in MP-EVOO and 13 in HP-EH-EVOO) followed an EEG resting-state recording session (eyes-open and closed) before and after the treatment. Following our dominant coupling mode model (DoCM), we built a dynamic integrated dynamic functional connectivity graph (iDFCG) that tabulates both the functional strength and the DoCM of every pair of brain areas.

**Results:** Signal spectrum within 1-13 Hz and theta/beta ratio have been decreased in the HP-EH-EVOO group in both conditions. ***FI***^***DoCM***^ has been improved after the intervention across groups and conditions but was more prominent in HP-EH-EVOO group (p < 0.001). Finally,we revealed a significant higher post-intervention reduction of NI (ΔNI^Total and α^) for the HP-EH-EVOO compared to the MP-EVOO and MeDi groups (p < 0.0001).

**Conclusions:** Long-term intervention with HP-EH-EVOO reduced the over-excitation of information flow in spontaneous brain activity.

**Significance:** Our study confirms the alteration of signal spectrum of EEG rhythms and dominant coupling mode due to the intervention with HP-EH-EVOO nutrition protocol.

**Highlights:**

- Non-pharmaceutical intervention based on HP-EH-EVOO in MCI reduces the over-excitation of information flow
- Non-pharmaceutical intervention based on HP-EH-EVOO in MCI increases the human brain flexibility
- Reconfiguration of dominant coupling modes in EEG resting-state due to the intervention is modulated by alpha frequency

## 1. Introduction

Mild cognitive impairment (MCI) is characterized as a multi-factorial syndrome that depends on psychological, neurobiological and social factors that all contributed to a high risk for developing dementia (Petersen et al., 1999). Clinical judgement of MCI demands multiple sessions and multiple domain tests to detect a cognitive decline and impairment across single or various cognitive functions (Winblad et al., 2004). Distinctions over multiple (amnestic or non-amnestic) and single domain MCI are more than important bearing the fact that amnestic-MCI single-domain, amnestic-MCI multiple-domain and mild Alzheimer’s disease (AD) could represent a continuum process that leads to deviations from normal aging (Brambati et al., 2009).

Besides the heterogeneity of pathophysiological mechanisms underlying MCI subjects and their diagnostic misidentification, there are no pharmaceutical therapeutic options available and approved by FDA or EMA. The pharmaceutical interventions include acetylcholinesterase-inhibitors and memantine which are approved only for AD and not for MCI and other medications which are not yet approved such as, ginkgo biloba, anti-inflammatory drugs, statins, platelet aggregation inhibitors (trifusal), piracetam and others (Karakaya et al., 2013). However, clinical trials based on the aforementioned substances targeting to neurofibrillary tangle formation and amyloid-beta accumulation failed to show a high efficient performance measures for AD patients with both primary and secondary cognitive parameters.

Recent evidences from non-pharmaceutical interventions in MCI and dementia subjects open new avenues over alternative options of treatments (Vlachos and Scarmeas, 2019). Recent studies reported promising evidences of alternative non-pharmaceutical treatments for MCI based on natural substances (e.g. Extra Virgin Olive Oil (EVOO) (Tsolaki et al 2020), saffron (Tsolaki et al., 2016)) and dietary interventions (e.g. the Mediterranean diet (MeDi)) (Andrich et al., 2015);. The MeDi is not a prescribed diet by a clinician but a general eating habituation from people located geographically around the Mediterranean sea. The traditional Mediterranean diet is characterized by high consumption of olive oil, vegetables, legumes, fruits and unprocessed cereals, moderate to high consumption of fish, low consumption of meat and meat products, and low to moderate consumption of dairy products. It is also characterized by moderate wine consumption. Scarmeas et al., reported that the adaptation of MeDi diet for a long period of time improved various cognitive functions and decreased the risk to develop MCI and also to progress from MCI to AD (Scarmeas et al., 2009). Moreover, healthy dietary habituations are strongly correlated with positive cognitive outcomes (Scarmeas et al., 2018).

Positive evidences of EVOO-enriched diet against AD pathology have been evaluated in both animals and humans. Researchers explored if the consumption of EVOO-enriched diet from TgSwDI mice could reduce the accumulation of amyloid- and tau-related brain levels and also the related alterations of cognitive functions. They reported that a long-term (6 months) consumption of EVOO containing diet provided a protective mechanism against AD pathology (Qosa et al., 2015). A clinical study investigated the potential positive role of EVOO-rich MeDi in 285 subjects with high vascular risk. The authors reported an improved cognitive functioning compared to a control diet. Additionally, participants followed EVOO-rich MeDi had a less number MCI subjects at follow up that the controls while participants followed a MeDi + Nuts didn’t differ from the controls in terms of the number of MCI subjects (Martinez-Lapiscina et al., 2013) after 7 years follow up.

The main ingredients of EVOO are glycerides of fatty acids (98%), mainly of mono-unsaturated fatty acids (MUFA) and especially of oleic acid. The remaining 2% include a variety of “minor compounds” like phenolic alcohols, phenolic acids, lignans, flavones and importantly secoiridoids and their derivatives (Servili et al., 2002, 2013). Natural phenols can modulate cell redox state (Singh et al., 2008) via a direct activity over enzymes, proteins, receptors and various signalling pathways (Kim et al., 2014; Goszcz et al.,2017). Another positive impact of natural phenols is their interference with biochemical homeostasis (Abuznait et al., 2013; Grossi et al., 2013) which further has impact to epigenetic alterations of the chromatin (Ayissi et al., 2014; Declerck et al., 2016).

There is strong evidence that MeDi prevents neurodegeneration due to the consumption of vegetables, legumes, and olive oil riched in polyphenols (Feart et al., 2010, 2013; Scarmeas et al., 2006, 2009, 2011). Phytochemicals which are chemical compounds produced by plants like flavonols, polyphenols support different protective biological activity of the cell. The repertoire of this activity could be anti-oxidant, anti-allergic, anti-inflammatory, anti-viral, anti-carcinogenic and anti-proliferative (Middleton, 1998; Hollman et al., 1999; Eastwood,1999). MeDi is characterized complementary to fruits and vegetables consumption by a consumption of EVOO in amounts ranging from 25 to 50 g/day (López-Miranda et al., 2008; Pitozzi et al., 2012; Qosa et al., 2015). Studies reported that polyphenols act as an anti-aggregation agent against the desposition of amyloid-β_1-42_ peptide plaques supporting an anti-toxic environment (Rigacci et al., 2011), while oleocanthal is capable of altering the oligomerization state of oligomers of amyloid-β_1-42_ peptide, which are toxic (Pitt et al., 2009). Both substances could have a great impact over the modifying treatment of AD.

A link between cognitive performance and dietary habits has been evaluated (Otaegui-Arrazola et al., 2014) while neurodegenerative disorders and cognitive decline have been associated to oxidative stress (von Bernhardi et al., 2012). A simple hypothesis from the aforementioned evidence of beneficial properties of antioxidant-rich foods is that their consumption could reduce the oxidative stress which could further protect from neurodegenerative diseases. MeDi is full of anti-oxidant rich foods with main origin from plants and has been associated with a health status (Sofi et al., 2010).

Normally, cells can counteract the oxidative insults by regulating their homeostasis. In neurodegenerative conditions related to aging, cell’s capacity to maintain a redox balance decreases which further has a consequence to mitochondrial dysfunction, neuronal injury and accumulation of radicals. We know that oxidative stress increases with aging (Finkel and Holbrook, 2000) and it is a significant age factor that makes neuronal systems more vulnerable to various neurodegenerative diseases (McCord, 2000). Oxidative stress causes mitochondrial injury which further causes loss of oxidative metabolism and an overproduction of free radicals. Oxidative stress makes a significant contribution in many human diseases such as AD (Lyras et al.,1999) and Parkinson disease (Cohen et al.,1984).

Our MICOIL Pilot Study was the first-ever reported longitudinal double-blind study which administered Greek HP-EH-EVOO to people with MCI for one year. The purpose of MICOIL was to evaluate the effect of a Greek HP-EH-EVOO+MeDi (Group 1) compared to a Greek MP-EVOO+MeDi (Group 2) and MeDi only (Group 3) on the multi cognitive status of patients with MCI, who are community-dwelling, Greek-speaking elderly, with age ranging from 60 to 80 (Tsolaki et al., 2020). The Greek HP-EH-EVOO received (50mL/day) together with MeDi instructions, the GreekMP-EVOO received (50mL/ day) together with MeDi instructions and MeDI group received only the MeDi instructions. The evaluated hypothesis that was that long-term intervention with either HP-EH-EVOO or MP-EVOO could be correlated with a significant improvement of cognitive functions compared to MeDi, independently of the presence of APOE ε4 carrier.

In the present study, we analysed electroencephalographic (EEG) resting-state recordings (both eyes-open and eyes-closed conditions) from a subset of participants from the three groups compared to the previous study focusing only on neuropsychological estimates (Tsolaki et al., 2020). EEG activity was recorded before and after the intervention with the three groups. We adopted a dynamic functional connectivity analytic pathway with the incorporation of our dominant coupling mode model (DoCM). Based on our DoCM, we incorporated both within frequency (intra-frequency) and between frequencies (cross-frequency) functional interactions under the same model. The main outcome of DoCM is the construction of an integrated dynamic functional connectivity graph (iDFCG) that tabulates both the functional strength and the DoCM of every pair of brain areas across experimental time (Dimitriadis, 2018). When two brain areas communicate under the same frequency content (intra-frequency) then this exchange of information is called linear. However, cross-frequency interactions are nonlinear information pathways and are highly active when two functionally distinct brain areas want to interact and exchange information (Hyafil et al., 2015). The ratio of cross-frequency over intra-frequency interactions can build a nonlinearity index (NI) of information change in our brain. In a resting-state EEG study, we showed in a group of adolescents with schizophrenia spectrum disorders (SSDs) that NI is significant higher compared to an age-matched healthy group and this reconfiguration of dominant coupling modes was driven by the alpha frequency (Dimitriadis, 2020). Here, we will adopt the same methodology assuming that the three groups will show a reduction of their individual NI after the treatment and also group1 and 2 administered with either Greek HP-EH-EVOO or Greek MP-EVOO will show a stronger reduction of their NI compared to the MeDi group 3. We hypothesized that the change of NI which is interpreted as a reduction of cross-frequency coupling (CFC) non-linear interactions will be driven by alpha frequency band which is a frequency of arousal at resting-state conditions (Barry et al., 2007). Eyes-open and eyes-closed are not two equivalent baseline ones and should be treated differently. Eyes-closed condition can be used as an arousal baseline while eyes-open condition as an activation baseline.

## 2. Material and Methods

### 2.1 MICOIL Study Design

From 1 December 2016 to 30 August 2018, participants were recruited from the Memory & Dementia clinic of the 3rd and 1st Neurology Departments of Aristotle University of Thessaloniki, Greece, and from the two Day Centers of the Greek Association of Alzheimer’s Disease and Related Disorders (GADRDA). The MICOIL study was realised in accordance with the Declaration of Helsinki and was approved by the GADRDA scientific & ethics committee (25/ 240 2016) (Clinical Trials Registration Number: NCT03 241 362996)^1^. All participants fulfilled the Petersen criteria for MCI (Petersen et al., 2009). Information about inclusion and exclusion criteria, study design, APOE genotype, randomization and allocation procedure of participants’ selection, withdrawals and description of every protocol followed for each group, we forward an interested reader to the first report of the MICOIL study (Tsolaki et al., 2020).

### 2.2 Participants

Demographics of the subjects that participated on EEG data collections is given in Table 1. Following a Kruskal-Wallis non-parametric test, we didn’t detect any effect over the groups in terms of the age, gender ratio and education level.

**Table 1.** Mean and standard deviation (SD) of demographics of all participants, and the results of the non-parametric Kruskal-Wallis statistical test

|  | **MeDi**<br>**(n=14)**<br>**mean±std** | **MP-EVOO**<br>**(n=16)**<br>**mean±std** | **HP-EH-**<br>**EVOO**<br>**(n=13)**<br>**mean±std** | **All**<br>**(n=43)**<br>**mean±std** | **Kruskal-**<br>**Wallis test**<br>**H(df)=x<sup>2</sup>,p** |
| --- | --- | --- | --- | --- | --- |
| **Age (Years)** | 70.5±5.9 | 71.2±7.9 | 69.5±7.6 | 70.5±7.1 | H(2) = 0.47<br>**p=0.78** |
| **Gender**<br>**(F:M)** | 12:2 | 11:5 | 7:6 | 30:13 | H(2) = 3.18,<br>**p = 0.20** |
| **Education**<br>**(Years)** | 10.9±4.7 | 9.1±3.6 | 11.4±3.0 | 11.4±3.0 | H(2) = 3.29,<br>**p = 0.19** |

### 2.3 Neuropsychological Assessment

The neuropsychological battery included the Greek version of Mini-Mental State Examination (MMSE) [27, 28] to assess the general cognitive function, Rivermead Behavioral Memory Test-Story Immediate and Delayed recall [29, 30] for episodic memory, Rey Osterrieth Complex Figure Test copy and delayed recall [31, 32] which measures visuospatial long-term memory and executive functions, Trail Making Test parts A & B [33], to examine visuospatial ability, attention and executive functions, Alzheimer Disease Assessment Scale-Cognition (ADAS-Cog) [34, 35] to assess the severity of cognitive dysfunction, Wechsler Memory Scales Digit Span Forward and Backward [36, 37] to assess attention and working memory, Letter and Category Fluency Test [38] for assessing phonemic and semantic fluency and Clock-drawing Test [39] which measures visuo-spatial orientation, understanding of verbal instructions, abstract thinking, planning, concentration, executive and visuo-spatial skills. Depressive symptoms were assessed by the Geriatric Depression Scale [40, 41] using a cut-off score of < 6 at baseline. We also used the Neuropsychiatric Inventory [42, 43] for the assessment of other neuropsychiatric symptoms, since it is a critical component for the evaluation of the MCI subjects because their distress can cause or exacerbate cognitive problems. For further details regarding the neuropsychological assessments, see the first paper focusing on neuropsychological estimates (Tsolaki et al., 2020).

### 2.4 EEG Recordings

EEG experiments are performed in cognitive intact elderly subjects and subjects with suspect of neurodegenerative disorder and cognitive deficits. To reduce the time of the EEG montage, we should use a minimum of 19 standard exploring electrodes placed according to basic international 10-20 system (i.e. Fp1, Fp2, F7, F3, Fz, F4, F8, T3, C3, Cz, C4, T4, T5, P3, Pz, P4, T6, O1, and O2). A higher number of EEG electrodes extending the basic 10-20 system is appreciated. For all EEG recordings, there was inclusion of two ear (A1 and A2). Resistance of the EEG electrodes should be lower than 5 KOhm.

EEG experiment included with the resting state EEG conditions:

1. Resting state eyes closed EEG recording for 5 minutes (ideally, most of these 5 minutes of EEG recordings should be characterized by a subject relaxed not showing voluntary or involuntary movements).
2. Resting state eyes open EEG recording for 5 minutes.

The sampling frequency was set to fs = 500 Hz.

EEG time series were re-referenced to the average reference electrode (Nunez and Srinivasan, 2006) before pre-processing steps. The program of experiments has been approved by the local ethical committee of Alzheimer Hellas (no. 25/21-6-2016).

### 2.5 Artifact reduction with independent component analysis (ICA) and Wavelet Decomposition

An important pre-processing step before the estimation of dynamic functional connectivity graph (dFCG) is the denoising of EEG recordings. We adopted an already established algorithmic artefact reduction method that combines independent component analysis (ICA) and Wavelet Decomposition. For further details, see our recent study which describes in detail the adopted denoising approach (Dimitriadis, 2020). Briefly, the majority of studies adopted ICA that produces N independent components (ICs) where N denotes the number of EEG sensors. Afterward, we rejected ICs as artifactual components based on the time course of ICs, the topology of the related weights per sensor and also estimating measures like entropy, kurtosis and skewness to further support our decision. The main drawback of this approach especially for resting-state is that we have to zeroing one IC as a whole even if any kind of artifact is detected in specific epochs covering a small percentage of the total experimental time. For that reason, we decided in our previous study to decompose the time course of artifactual IC in epochs of 1 sec using wavelet decomposition. This approach gives us an advantage over traditional approach avoiding rejecting an IC that encapsulates also critical true brain activity mixed with a number of artifactual epochs (Dimitriadis, 2020).

### 2.6 Signal Power Analysis

We estimated power spectral density (PSD) using *pwelch* MATLAB function independently for every EEG sensor for each subject before and after the intervention and in both conditions. Then, we averaged PSD estimations across EEG sensors to characterize every subject’s PSD profile for every condition and in both pre and post intervention period. The PSD analysis was realized up to 45 Hz. We also estimated the frontal theta/beta ratio as a marker of attentional control (Putman et al., 2014; Clarke et al.,2019).

### 2.7 The integrated dynamic functional connectivity graph (IDFCG) based on iPLV

In this section, we will describe briefly our dominant intrinsic coupling model (DoCM) demonstrated in the whole repertoire of functional neuroimaging modalities (Antonakakis et al., 2017; Dimitriadis et al., 2017a-c, 2018a-d, 2019; Dimitriadis, 2020; Marimpis et al., 2020). The majority of studies so far were investigating functional connectivity independently for every frequency band while they focused mainly on within-frequency (intra-frequency) interactions. Last years, an increased amount of research studies exploring also functional connectivity between brain areas oscillating on a different frequency the so-called cross-frequency coupling (CFC) or inter-frequency interactions. DoCM model incorporates all possible functional connectivity modes under a same framework. The main aim of DoCM model is to detect the dominant coupling mode between every pair of EEG sensors and across temporal segments. Here, we adopted intra-frequency phase-to-phase and inter-frequency phase-to-amplitude potential coupling modes (CFC). We estimated dynamic functional connectivity graph (dFCG) within and between the seven studying frequency bands {δ, θ, α1, α2, β1, β2, γ} defined, respectively, within the ranges {0.5–4 Hz; 4–8 Hz; 8–10 Hz; 10–13 Hz; 13–20 Hz; 20–30 Hz; 30–48 Hz}. For this computation, we employed the EEG activity from the 19 EEG sensors in both resting-state conditions. EEG recordings were bandpass filtered using a 3^rd^ order zero-phase Butterworth filter employing the *filtfilt* MATLAB function. The width of the temporal window was set equal to 500ms (or 250 samples) and moved forward across experimental time with a step equals to 100 ms (50 samples) which can encapsulate both slow and fast oscillations (Dimitriadis et al., 2013a,b, 2015a,b, 2016a,b,c, 2017a,b, 2018b,c; Marimpis et al., 2020). We finally analyzed 75 secs across subjects and conditions as a common experimental time across the participants.

For every pair of EEG sensors and for every temporal segment, we estimated the seven intra-frequency phase-to-phase possible interactions, one for every frequency band and twenty-one inter-frequency phase-to-amplitude interactions between every possible pair of the seven frequency bands. Here, we adopted iPLV as the proper connectivity estimator for both intra and inter-frequency coupling modes as in our previous studies. In total, we estimated twenty-eight possible coupling modes leading to twenty eight dFCG per subject and condition. A dFCG is a 3D matrix of dimensions [temporal segments x sensors x sensors]. Then, we followed a surrogate analysis with main scope to detect the dominant coupling mode per pair of EEG sensors and for each temporal segment (for further details see Dimitriads, 2020). This approach leads to the integrated dFCG (DIFCG) that keeps both the strength and also the dominant coupling mode per pair of EEG sensors and across experimental time. The IDFCG is a pair of 3D matrices of size [temporal segments x sensors x sensors] where one keeps the strength and the other the dominant coupling mode. The strength is a value of range [0,1] based on the adopted connectivity estimator iPLV while the dominant coupling mode is encoded with an integer from 1 up to 28 e.g 1 for δ, 2 for θ, 3 for α_1_, …, 26 for β_1_-β_2_, 27 for β_1_-γ and 28 for β_2_ - γ.

Fig. 1 is created in analogy to previous studies to further exemplify the concept of DoCM model in not familiar readers (Dimitriadis, 2020; Marimpis et al., 2020). Fig.1A illustrates how DoCM is defined for the first two temporal segment of eyes-closed condition from the first subject of group A between Fp1 and Fp2 EEG sensors. Following the important step of surrogate analysis, we revealed α_1_-β_2_ cross-frequency phase-to-amplitude coupling (PAC) as the dominant coupling mode (DoCM) among 28 possible coupling modes for the first two temporal segments for this particular EEG pair of sensors. On the right side of frequency-dependent pairs of time series, we demonstrated as a matrix the functional strength measured with iPLV OF every possible coupling mode. The main diagonal of this matrix tabulates the intra-frequency phase-to-phase coupling modes while the off-diagonal stores the cross-frequency phase-to-amplitude coupling modes. Fig.1B illustrates the temporal evolution of DoCM for the Fp1-Fp2 EEG pair. The color encodes the functional strength of the coupling while the y-axis refer to the detected DoCM. On the right side of this semantic time series, we showed the probability distribution (PD) of DoCM across experimental time. From this representation, α_1_-α_1_ intra-frequency phase-to-phase coupling was the most representative across DoCM. Both the flexibility Index (FI) and PD are estimated from semantic time series as the one demonstrated in Fig. 1B.

**Figure 1.**
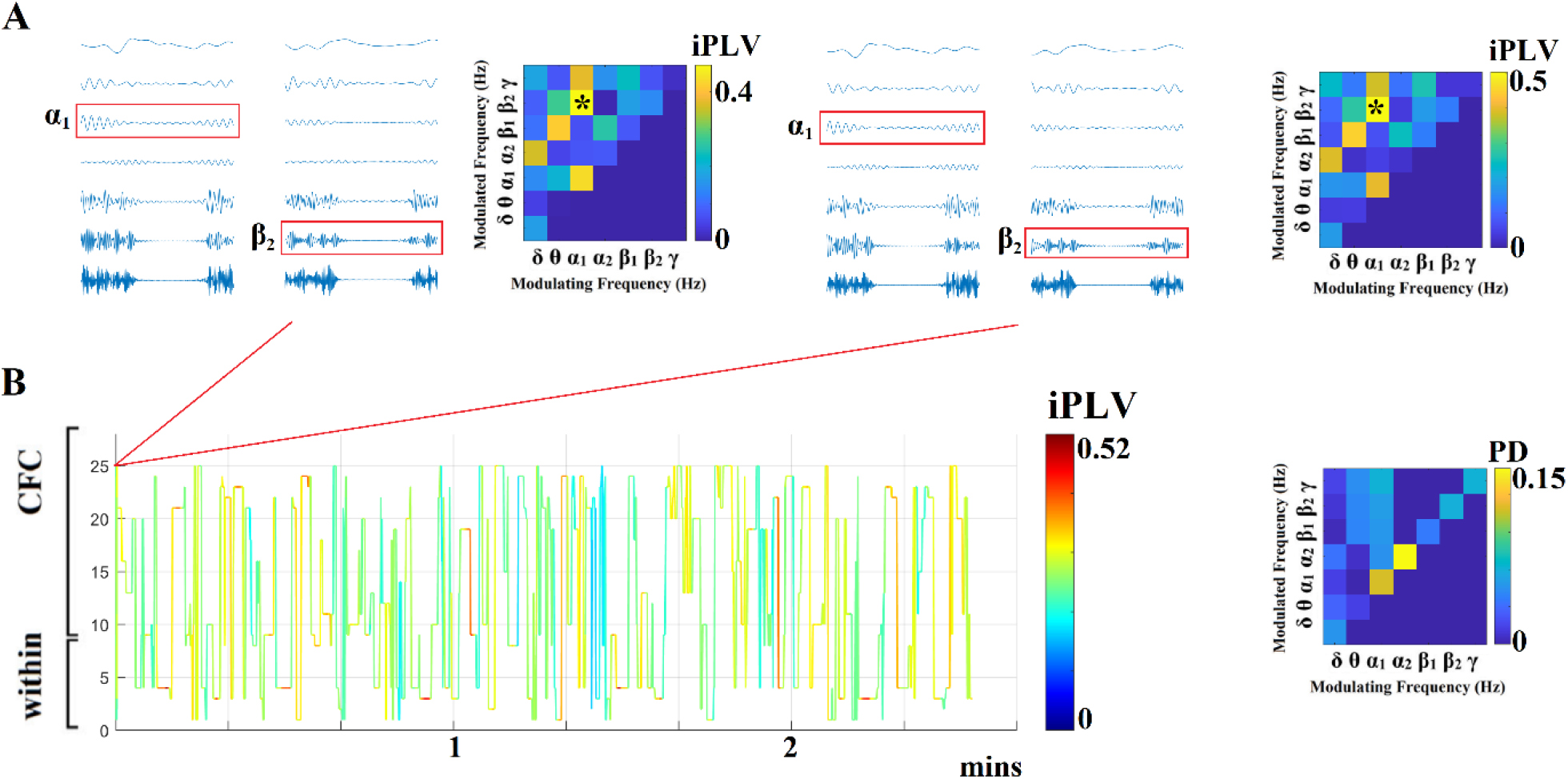
Detecting Dominant Intrinsic Coupling Modes (DoCM) based on iPLV. (A) A schematic illustration of the adapted DoCM model showing the detection of dominant coupling mode between EEG sensors Fp1 and Fp2 for the first two consecutive temporal segment (ts_1_, ts_2_). Surrogate analysis revealed the DoCM for both temporal segments. During both ts1 and ts2, the DoCM reflected significant phase-to-amplitude coupling between α_1_ phase and β_2_ amplitude (indicated by red rectangles). Both colored matrices tabulate the functional strength quantified with iPLV of both intra-frequency coupling modes (main diagonal) and inter-frequency coupling modes (off-diagonal). (B) Burst of DoCM between the Fp1 and Fp2 sensors. The colored time series is a streaming of information flow between two brain areas where the main elements of this neural communication are the DoCM. This streaming of DoCM can be interepreted as a temporal messaging of brain areas in the macroscale level (Buzsáki and Watson, 2012). The y-axis in B. encodes the DoCM dividing the axis into intra and inter-frequency coupling modes. The matrix on the right tabulates the probability distribution (PD) of DoCM for the semantic time series showed on the left. α1-α1 phase-to-phase intra-frequency coupling was the most prominent among 28 potential coupling modes. The flexibility index (FI) is estimated over a semantic time series showed in B. by quantifying the number of times a DoCM changed between consecutive temporal segments divided by the number of temporal segments – 1. FI ranges within [0,1] where high values can be interpreted as functionally more flexible communication between brain areas.

### 2.8 Semantic features derived from the evolution of DoCM

This section describes in detail the semantic features that can be extracted from the 2^nd^ 3D tensor that preserves the DoCM across EEG sensor space and experimental time.

#### 2.8.1. Flexibility index (FI)

We estimated Flexibility index (FI), a measure that quantifies the transition rate of DoCM between every pair of EEG sensors (Dimitriadis and Salis, 2017; Dimitriadis et al., 2018d; Dimitriadis, 2020). We employed the 2^nd^ 3D tensor of the DIFCG that tabulates the semantic information of DoCM across the brain and experimental time to estimate FI. An example of what information this tensor tabulates is shown in Fig.1B.

This metric will be called hereafter FI^DoCM^ and it is defined as:

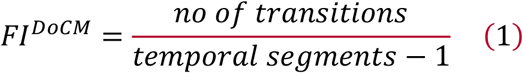

We estimated the *ΔFI*^*DoCM*^ between pre and post intervention *FI*^*DoCM*^

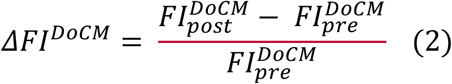

We counted a transition only when a coupling exist between two consecutive temporal segments. The term (temporal segments – 1) equals the pairs of temporal segments where a functional coupling mode exists. FI measure counts only the change of the DoCM across time and not the preferred transition between specific pairs of frequency coupling modes. FI^DoCM^ gets higher values for higher “transitions” of DoCM between neighboring in time temporal segments. Fig. 1B illustrates how FI^DoCM^ is estimated for the Fp1-Fp2 EEG pair. The outcome of FI estimation for every pair of EEG sensors is a matrix of size 19 × 19. Then, we estimated the nodal FI as the mean of every row of this matrix. Finally, we estimated the global FI by averaging the 19 nodal FI.

#### 2.8.2. Spatiotemporal distribution of DoCM—Comodulograms

Based on the 2^nd^ 3D DIFCG that keeps the semantic information of the preferred dominant coupling mode, we can tabulate in a frequencies × frequencies matrix the probability distribution (PD) of observing each of the DoCM frequencies across 7 (intra-frequency) + 21 (crossfrequency coupling) = 28 possible coupling modes. We estimated PD across sensor space between the EEG sensor pairs and also across temporal segments.

The spatiotemporal PD tabulated in a matrix is called hereafter comodulogram and an example is demonstrated in Fig. 1B (Antonakakis et al., 2016, 2017a, 2017b; Dimitriadis and Salis, 2017; Dimitriadis et al., 2018a, 2018b, 2018c).

#### 2.8.3. Nonlinearity index (NI) based on DoCM

When two brain areas communicates within the same frequency (intra-frequency) then this communication is linear. However, when two brain areas communicate via cross-frequency coupling then this pathway is called nonlinear and plays a pivotal important role in inter-areal communication (Chen et al., 2009, 2010; Dimitriadis, 2020). The distinction between within-frequency coupling (linear coupling) and cross-frequency coupling (non-linear coupling) has been validated via biophysical modeling (Chen et al., 2009, 2010). Nonlinear information pathways are highly active when two brain areas want to exchange information (Hyafil et al., 2015).

As it was mentioned in section 2.7.2, we estimated the PD of DoCM across EEG sensor space and at every temporal segment. The PD is divided in two sections, the intra-frequency interactions (7 in total) which are tabulated in the main diagonal in the comodulogram (Fig.1B) and the cross-frequency interactions in the off diagonal (21 in total).

The ratio of the sum of this 21 cross-frequency PD values versus the 7 intra-frequency values define our nonlinearity index (NI) (Dimitriadis, 2020) which is described in eq. 2. In our study as in a previous one (Dimitriadis, 2020), we assumed that any improvement (reduction) of NI after the intervention should be leaded by the α frequency due to the resting-state condition. To quantify the driving role of α frequency, we estimated NI as the ratio of the sum of PDs between α1 and {α_2_,β_1_,β_2_,γ} (4 PDs related to 4 cross-frequency coupling pairs) and between α_2_ and {β_1_,β_2_,γ} (3 PDs related to 3 cross-frequency coupling pairs) with the sum of PDs related to α_1_ and α_2_ within frequencies interactions (2 PDs related to 2 intra-frequency couplings) (eq. 3). The higher the NI the higher is the contribution of CFC to the DoCM and so the higher is the nonlinear communication between brain areas. The outcome of this process is a time series of size equal to the number of temporal segments which is hereafter will be called dynamic NI (dNI). Two dNI were estimated per subject and condition, one incorporating the whole repertoire of coupling modes and one targeting to the α frequency as a modulating frequency. The eq.3 and 4 report the estimation of NI^Total^ and NI^α^ for one temporal segment.

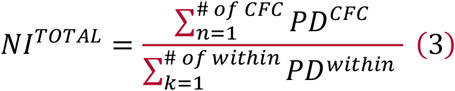

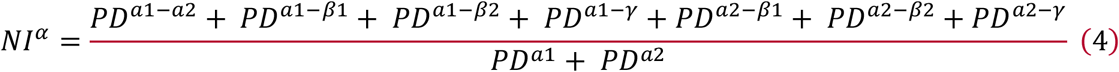

To quantify the level of potential difference of dNI between the pre and post-condition, we estimated the delta difference Δ of the median values of dNI.

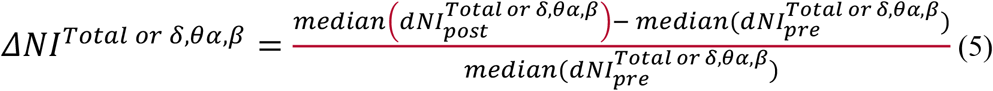

### 2.9 Statistical Analysis

We estimated Cohen’s d effect size for PSD as a mean across frequency bin between pre and post-intervention period per group and in both conditions. A Wilcoxon Signed Ranks Test has been applied to the theta/beta ratio between pre and post intervention period per group. We also performed statistical analysis between pairs of groups for detecting group differences in terms of ΔNI. We also applied statistical analysis over PD of DoCM independently per group between pre and post intervention condition to uncover reorganization of DoCM due to intervention. For both statistical comparisons over ΔNI and PD of DoCM, we adopted the one-way ANOVA with p < 0.05 corrected for multiple comparison.

We estimated the delta (Δ) difference of each neuropsychological assessment as for FI and NI with the following formula (post - pre)/pre. ΔNI^Total or α^ from the eyes-open condition were used as response variable and neuropsychological assessments as predictors following a multi-linear regression analysis. Clock Drawing, clock copy, digit span forward and digit span backward were excluded from the analysis due to many stable findings in pre and post intervention period (Δ = 0). Our analysis has been realized solely for the eyes-open condition due to the fact that neuropsychological assessments have been done with eyes-open. For that reason, eyes-open condition is more appropriate than eyes-closed.

### 2.10 Power Analysis

We estimated effect size and actual power analysis a posteriori on the estimated measurements of global ΔFI^Total^ and ΔNI^Total or α^. As a statistical test, we employed one-way ANOVA omnibus with fixed effects. Power (1 - b err prob) was set to 0.95 and α err prob to 0.05. We repeated the analysis independently for the four conditions ({eyes-open – eyes-closed}x{ΔNI^Total^,ΔNI^α^}).

### 2.11 Software

The analysis has been realized in the MATLAB environment (v2019b) using the signal processing toolbox. Fast ICA has been adopted from the fieldtrip toolbox. Dynamic functional connectivity has been based on in-house software provided on our github website: https://github.com/stdimitr/time_varying_PAC and https://github.com/stdimitr/docm_model

## 3. Results

### 3.1 Alterations of PSD after the intervention period across groups and conditions

Fig.2A-C illustrates the group averaged PSD for eyes-closed condition across the three groups and similarly Fig.2D-F demonstrates the group averaged PSD for eyes-open condition. We didn’t detect any significant difference between pre and post intervention periods across groups and conditions. However, we observed important trends where PSD was reduced across the studying spectrum (up to 45 Hz) mostly in HP-EH-EVOO and MP-EVOO groups.

**Figure 2.**
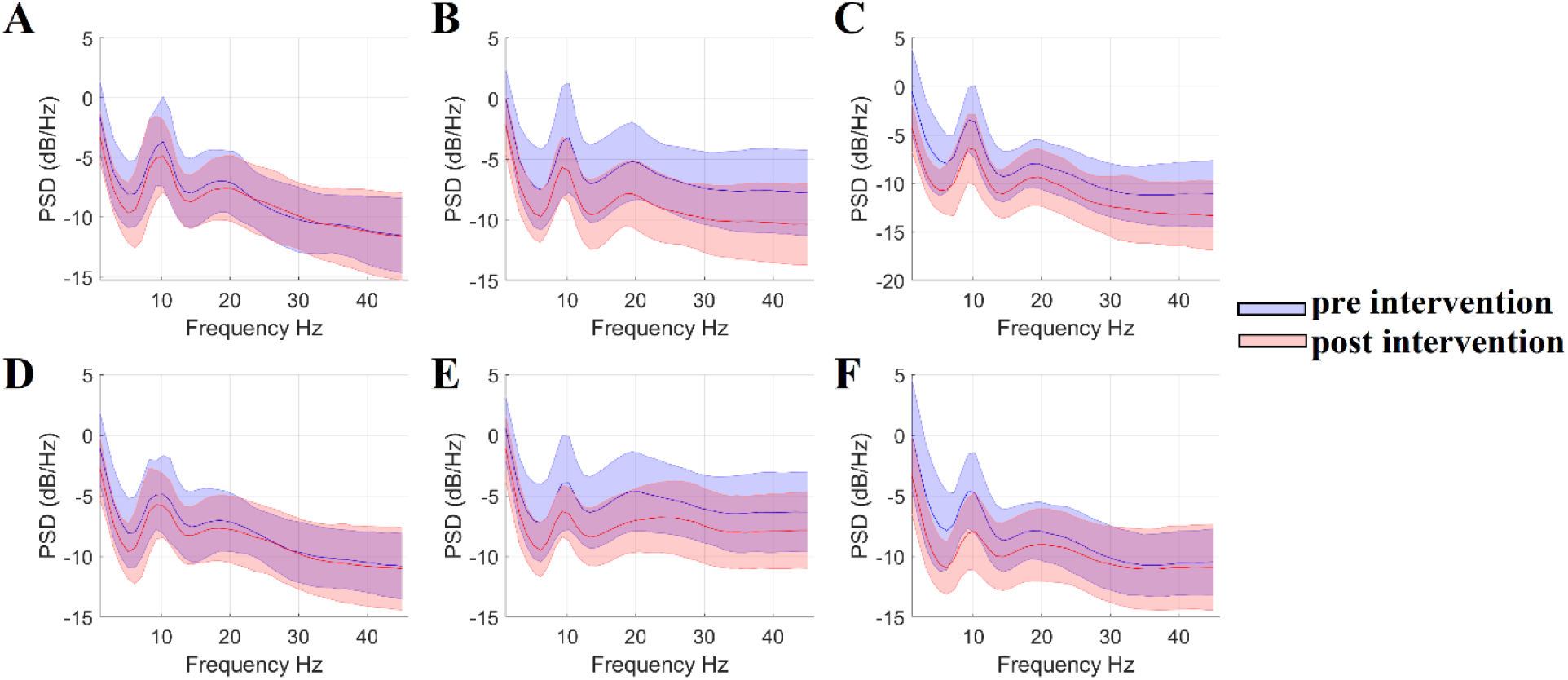
Group-averaged PSD across groups and conditions. A. Group-averaged PSD for MeDi in eyes-closed condition B. Group-averaged PSD for MP-EVOO in eyes-closed condition C. Group-averaged PSD for HP-EH-EVOO in eyes-closed condition D. Group-averaged PSD for MeDi in eyes-open condition E. Group-averaged PSD for MP-EVOO in eyes-open condition F. Group-averaged PSD for HP-EH-EVOO in eyes-open condition

**Figure 3.**
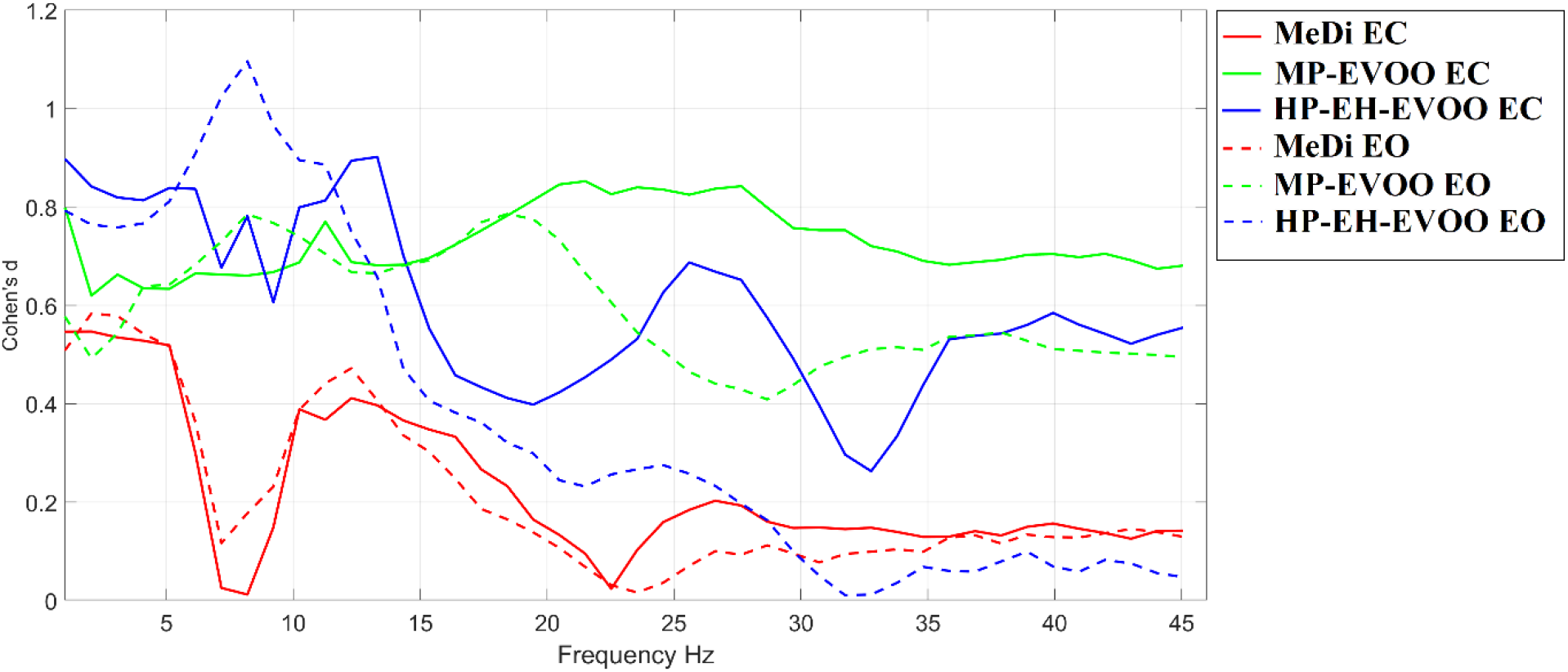
Cohen’s d effect size within the studying frequency spectrum forevery group and in both conditions. **(EC – Eyes Closed; EO – Eyes Open)**

In both conditions, we can clearly see large effect size for the HP-EH-EVOO, medium to large for MP-EVOO and small to medium for MeDi within 1 - 13 Hz (Table 2). A large effect size in beta band has been observed for the MP-EVOO group, and a small to medium effect for the HP-EH-EVOO and MeDi group. Table 2 summarizes the mean and standard deviation of the Cohen’s d effect size across frequency bins within specific frequency bands, the delta, theta, alpha and beta frequencies.

**Table 2.** Cohen’s d effect size of PSD across groups and in both conditions. Mean and standard deviations were estimated within specific frequency bands and across frequency bins. We underlined with bold the maximum effect size per frequency band across groups.

|  | Eyes - open |  |  |  | Eyes - closed |  |  |  |
| --- | --- | --- | --- | --- | --- | --- | --- | --- |
|  | 1-4 Hz | 5-8 Hz | 9-13 Hz | 14-30 Hz | 1-4 Hz | 5-8 Hz | 9-13 Hz | 14-30 Hz |
| **MeDi** | 0.55 ± 0.03 | 0.29 ± 0.18 | 0.38 ± 0.09 | 0.12 ± 0.09 | 0.53 ± 0.01 | 0.21 ± 0.24 | 0.34 ± 0.10 | 0.19 ± 0.09 |
| **MP-EVOO** | 0.56 ± 0.06 | 0.70 ± 0.06 | 0.70 ± 0.04 | **0.59 ± 0.13** | 0.67 ± 0.08 | 0.65 ± 0.01 | 0.69 ± 0.04 | **0.79 ± 0.05** |
| **HP-EH-EVOO** | **0.77 ± 0.01** | **0.96 ± 0.12** | **0.83 ± 0.12** | 0.26 ± 0.10 | **0.84 ± 0.03** | **0.78 ± 0.07** | **0.80 ± 0.11** | 0.52 ± 0.10 |

Interestingly, the frontal theta/beta ratio was significantly different only in HP-EH-EVOO in the eyes-open condition (p = 0.0032, zval = -2.7255; eyes – closed: p = 0.06, zval = -1.53). The theta/beta ratio was reduced in both conditions in HP-EH-EVOO group.

### 3.3 Improved *ΔFI*^*DoCM*^ for all the Groups

***ΔFI***^***DoCM***^ has been improved in the three groups in both conditions. However, the improvements are more prominent in HP-EH-EVOO and MP-EVOO groups (Fig.4).We didn’t observe any difference of the level of ***ΔFI***^***DoCM***^ between the two conditions. However, there is a trend of higher ***ΔFI***^***DoCM***^ **values for** HP-EH-EVOO and MP-EVOO in eyes-open compared to eyes-closed condition. Mean and standard deviations (st.d.) of the ***Δ****FI*^*DoCM*^ are tabulated in Table 3.

**Figure 4.**
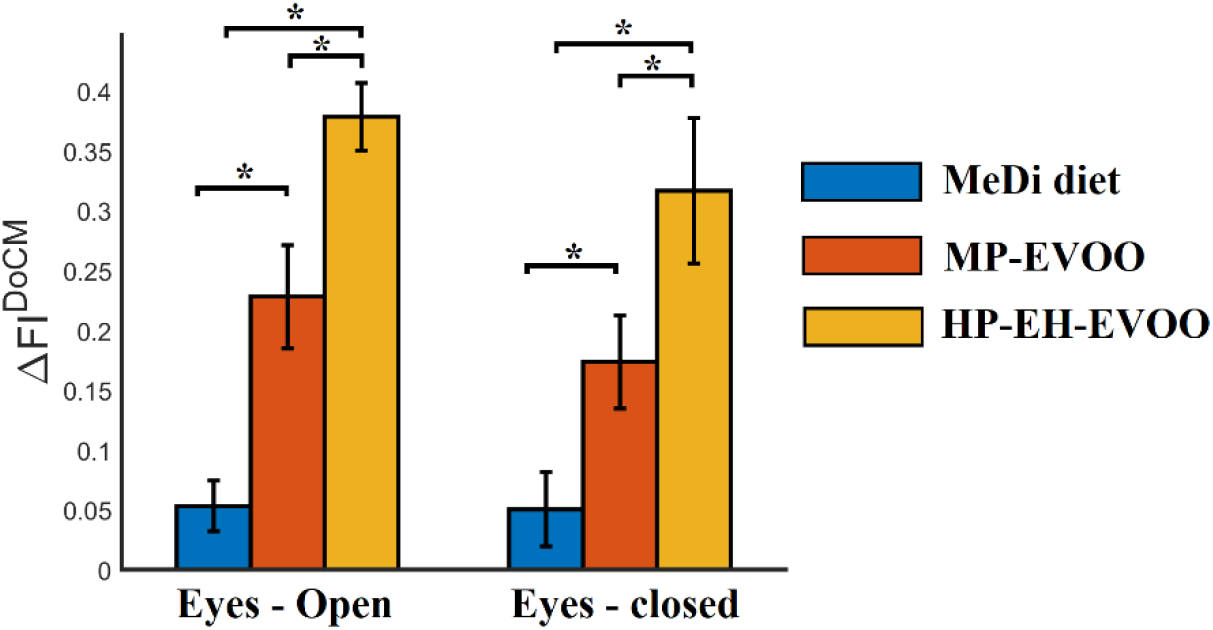
Group-averaged *ΔFI*^*DoCM*^ in eyes-open and eyes-closed resting-state conditions (* p < 0.0001, ANOVA one-way test)

**Table 3.** Mean and standard deviations of ***ΔFI***^***DoCM***^ in both groups.

|  | MeDi (mean/st.d.) | MP-EVOO<br>(mean/st.d.) | HP-EH-EVOO<br>(mean/st.d.) |
| --- | --- | --- | --- |
| **Eyes-open** | **0.05 ± 0.01** | **0.22 ± 0.01** | **0.37 ± 0.02** |
| **Eyes-closed** | **0.05 ± 0.01** | **0.16 ± 0.01** | **0.30 ± 0.02** |

### 3.4 ΔNI Group Differences Across Resting-State Conditions

Group-averaged dNI^Total^ and group-averaged dNI^α^ for the three groups and the two conditions are illustrated in Fig.5 and 6, correspondingly. Even though spontaneous activity is completely different across subjects, we reported group-averaged time-series in order to inform interested readers about the fluctuation of NI over time. Finally, we reported group-averaged ΔNI^Total or α^ in Fig.7 where there are significant differences between groups in both conditions and in both ΔNI^Total or α^. HP-EH-EVOO group showed significant higher post-intervention reduction of the NI level in total and in α in both conditions. Interestingly, group-averaged ΔNI^α^ were higher for eyes-open condition compared to eyes-closed in the three groups. Complementary, we observed interesting patterns for the rest of potential modulating frequencies. ΔNI^δ^ was increased after the intervention in both conditions and across groups (Fig.8A,D) while ΔNI^θ,β^ were reduced after the intervention in both conditions and across groups (Fig.8B,C,E,F). We detected the following significant differences between groups : i) the increment of ΔNI^δ^ was higher for MeDi compared to HP-EH-EVOO group in both conditions (Fig.8A,D) and ii) the reduction of ΔNI^β^ was higher for HP-EH-EVOO group compared to MeDi group in eyes-closed condition (Fig.8C).

**Figure 5.**
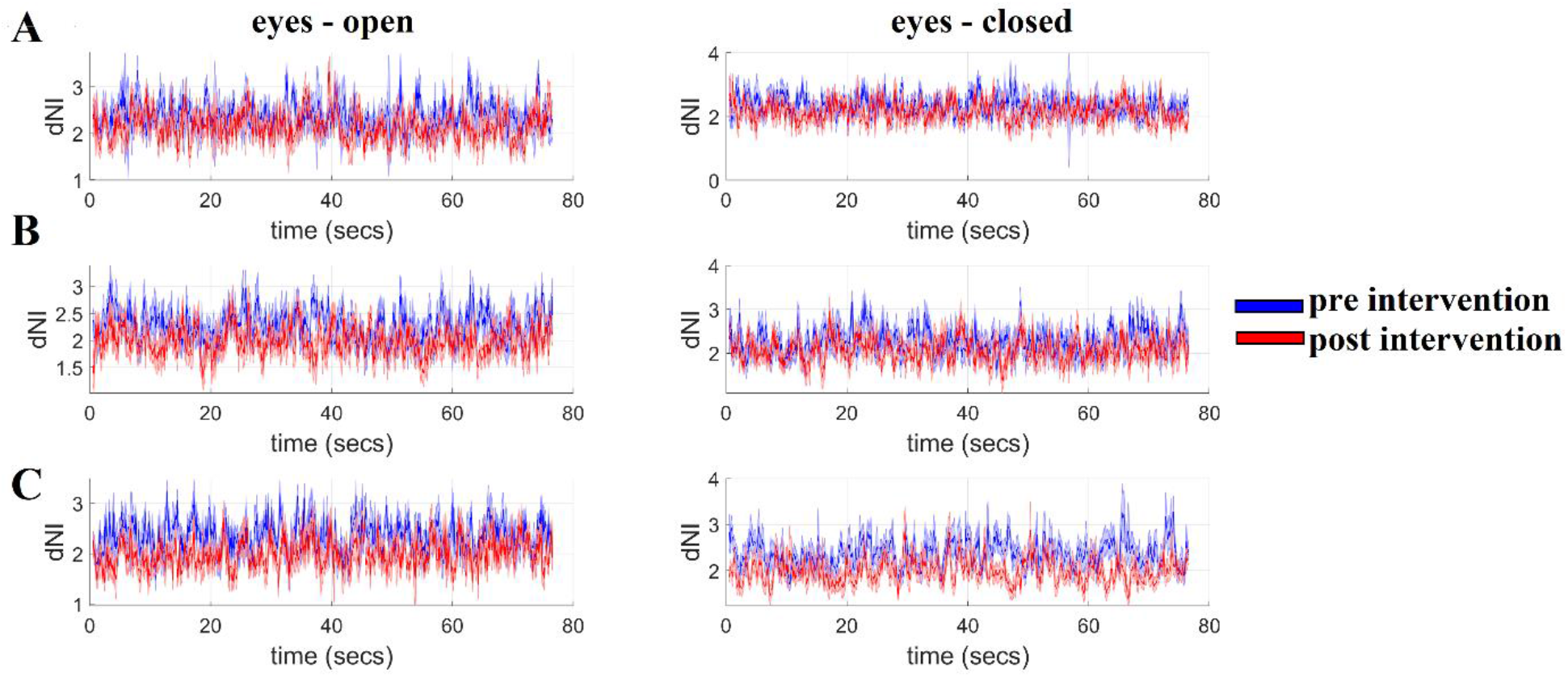
Group-averaged dNI per group and condition. **A**. MeDi diet group A **B**. HP-EH-EVOO group B **C**. MP-EVOO group C

**Figure 6.**
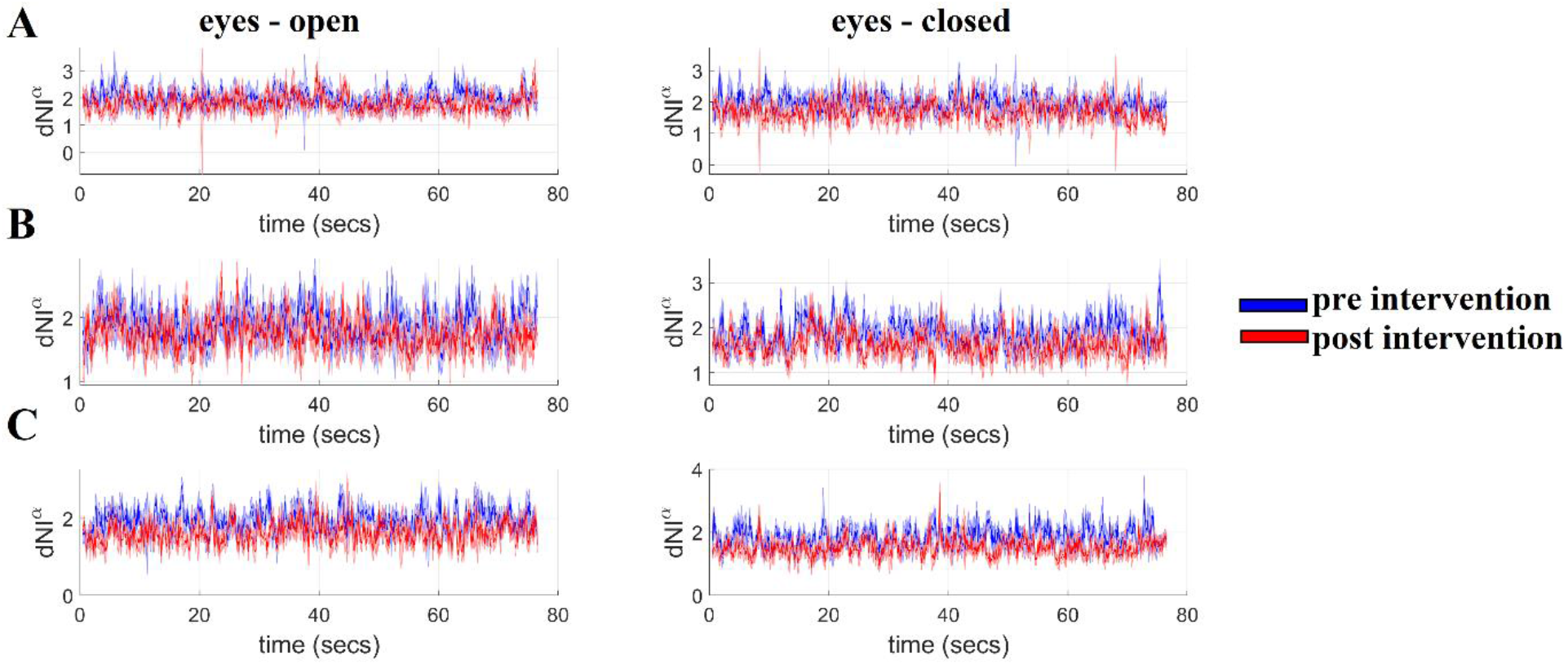
Group-averaged dNI^α^ per group and condition. **A**. MeDi diet group A **B**. HP-EH-EVOO group B **C**. MP-EVOO group C

**Figure 7.**
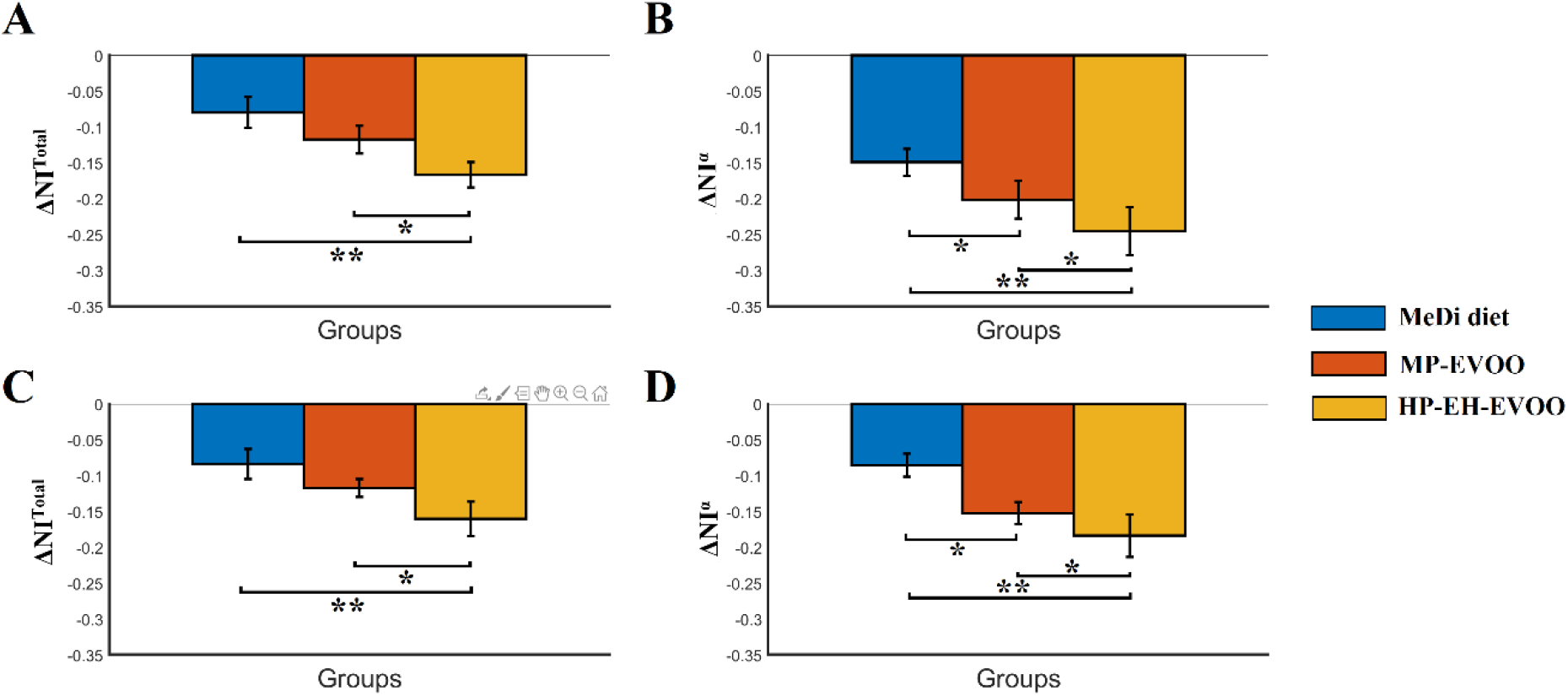
Group-averaged ΔNI^Total^ and ΔNI^α^ in both conditions. **A**. ΔNI^Total^ in eyes-closed condition **B**. ΔNI^α^ in eyes-closed condition **C**. ΔNI^Total^ in eyes-open condition **D**. ΔNI^α^ in eyes-open condition (* p < 0.001, ** p < 0.0001)

**Figure 8.**
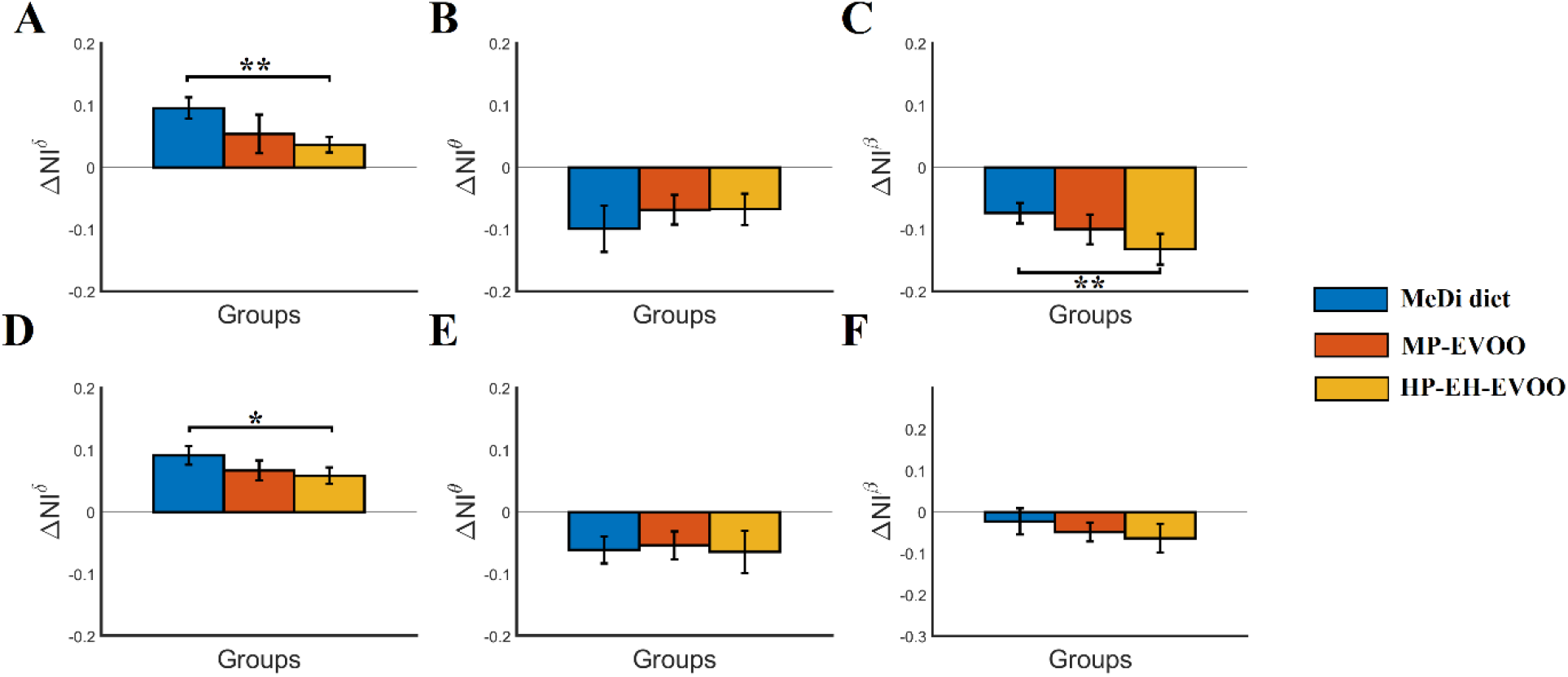
Group-averaged ΔNI^δ,θ,β^ in both conditions. A. ΔNI^δ^ in eyes-closed condition B. ΔNI^θ^ in eyes-closed condition C. ΔNI^β^ in eyes-closed condition D. ΔNI^δ^ in eyes-open condition E. ΔNI^θ^ in eyes-open condition F. ΔNI^β^ in eyes-open condition (* p < 0.001, ** p < 0.0001)

### 3.3 Power Analysis estimated over ΔFI and ΔNI findings

Table 4 reports the power analysis and the effect size of our ΔFI and ΔNI findings. It is clearly that our observations and statistical test are well powered by the sample size.

**Table 4.** Power analysis over ΔFI and ΔNI findings.

| | $\Delta FI$ | $\Delta NI^{Total}$ | $\Delta NI^{\alpha}$ |
| --- | --- | --- | --- |
| **Eyes-open** | P=0.99, f=4.23 | P=0.98 , f=3.56 | P=0.99, f=2.46 |
| **Eyes-closed** | P=0.99, f=2.47 | P=0.98 , f=2.00 | P=0.98, f=2.04 |

### 3.5 Multi-linear Regression Analysis

Multi-linear regression analysis for ΔNI^Total or α^ responses with the Δ differences of eight neuropsychological assessments as potential predictors revealed interesting findings. Below,we reported the R^2^,F-stattisti and the relevant pvalue for the two multilinear regression analyses:

i. R^2^=0.1758 F=2.4620 pvalue= 0.0365 for ΔNI^Total^
ii. R^2^=0.1217 F=1.8574 pvalue = 0.1070 for ΔNI^α^

β coefficients of both multi-linear regression analyses are illustrated in Fig.9. Our analysis untangled an interesting finding linking the reduction of ΔNI^Total^ with Δ differences of eight neuropsychological variables between the baseline and the follow up periods. Similar analysis with ΔNI^α^ didn’t reach the statistical level. However, the trend of β coefficients was similar with ΔNI^Total^ (Fig.9).

**Figure 9.**
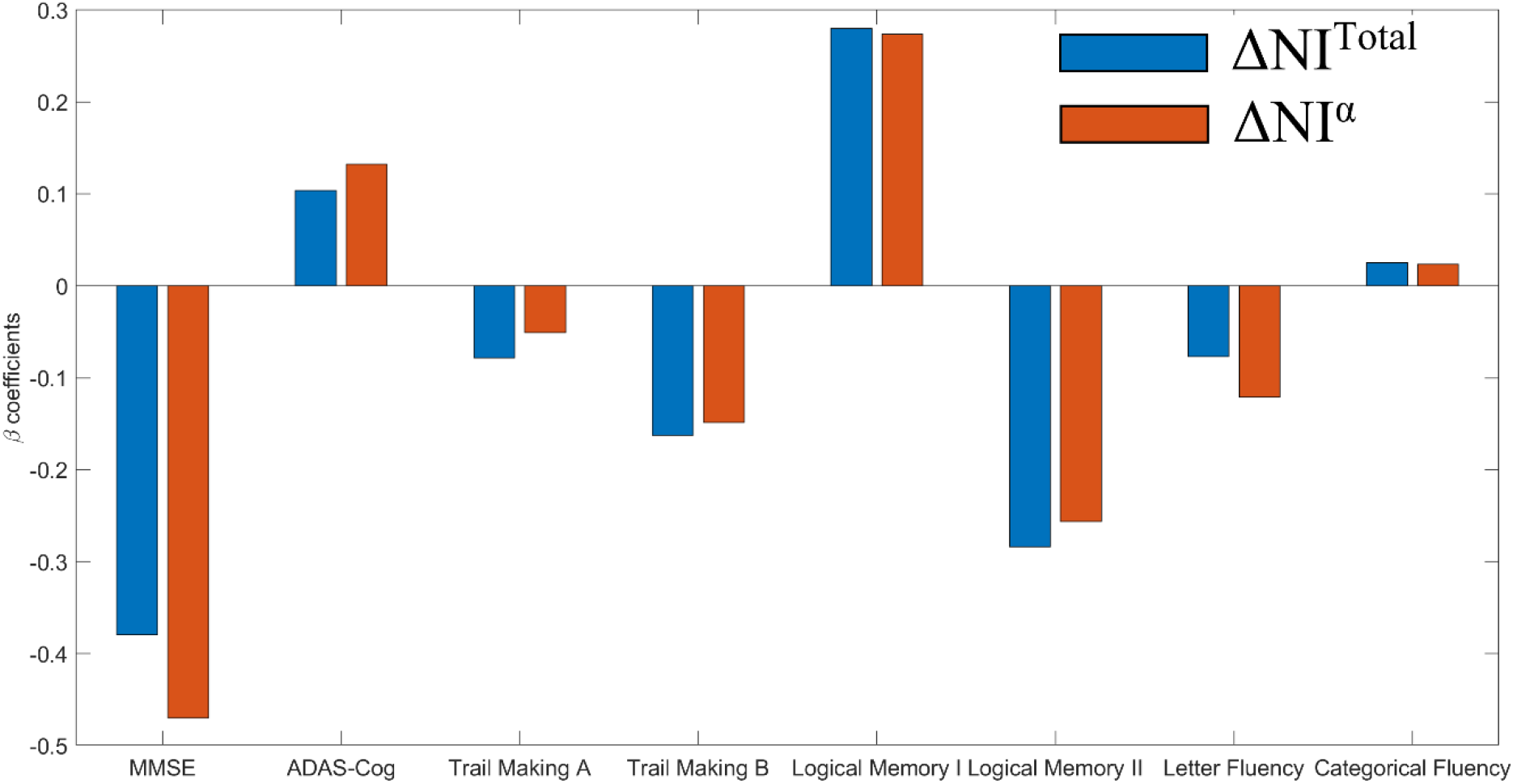
β coefficients of the multi-linear regression analysis for ΔNI^Total or α^ responses with the Δ differences of eight neuropsychological assessments as potential predictors.

## 4. Discussion

The present study is the first investigation of the effect of HP-EH-EVOO versus MP-EVOO and MeDi on the EEG brain dynamics at resting-state of people with MCI. In our previous study, we reported for the very first time that a long-term intervention with HP-EH-EVOO or MP-EVOO was associated with significant improvement in cognitive function compared to the MeDi group (Tsolaki et al., 2020). We adopted here our DoCM model which incorporates into a dFCG both the dominant coupling mode and also its functional strength between every pair of EEG sensros and acros experimental time (Antonakakis et al., 2017; Dimitriadis et al., 2017a-c, 2018a-d, 2019; Dimitriadis, 2020; Marimpis et al., 2020). We detected a significant higher post-intervention reduction of NI (ΔNI^Total and α^) for the HP-EH-EVOO compared to the MP-EVOO and MeDi groups. Our results support the reduction of the over-excitation of information flow across groups with a higher impact for the HP-EH-EVOO group.

Signal spectrum analysis didn’t reveal any significant difference between pre and post intervention periods across groups and conditions. However, we untangled an important trend By estimating the effect size within basic brain rhyhms (delta, theta, alpha and beta). In both conditions, we have observed a large effect size for the HP-EH-EVOO, medium to large for MP-EVOO and small to medium for MeDi within 1-13 Hz. A large effect size in beta band has been observed for the MP-EVOO group, and a small to medium effect for the HP-EH-EVOO and MeDi group.

Delta waves originate in medial frontal cortical areas and also in the insula an area associated with signals from our body (Mesulam and Mufson, 1982), the nucleus accumbens, and also the tegmental brainstem area” (Knyazev, 2012). Delta frequency was linked to an internal inhibition when it is important to inhibit all the irrelevant information from other sources. Delta frequencies up to 4 Hz originated in the frontal and anterior cingulate cortex have been linked to the inhibition of distracting information supporting the attentional demands of a task (Harmony, 2013). Global delta synchrony is suggested to be the result of GABAergic project neurons originating from the thalamic reticular nucleus (Herrera et al., 2019). The lateral geniculate nucleus, a second thalamic nucleus with a baseline attentional demands like in the eyes-open condition has been linked to the reduction of the global delta synchrony (Weynard et al., 2001). In our study, we observed a reduction of delta signal spectrum in both conditions mostly for the HP-EH-EVOO and secondly for the MP-EVOO underlined by large and medium effect size, correspondingly. Complemetary, ΔNI^δ^ was increased after the intervention in both conditions and across groups while the increment of ΔNI^δ^ was higher for MeDi compared to HP-EH-EVOO group in both conditions.

It is well-known that theta synchrony is involved in working memory processes (Sauseng et al., 2010). A recent study compared EEG signal power between mind wandering and cognitive tasks. They found an increased theta and a reduced beta power during mind wandering compared to cognitive tasks in a healthy group (Van Son et al., 2019). Especially, the defined frontal theta/beta ratio was increased and varied during mind wandering reflecting a reduced top-down attentional control favoring thoughts. Another study, reported a reduced frontal theta/beta ratio in healthy controls compared to a group with cognitive performance anxiety (Putman et al., 2014). The increased frontal theta/beta ratio is an index of top-down failure of attentional control. Here, we observed a significant reduction of frontal theta/beta ratio only for HP-EH-EVOO group in eyes-open condition. This reflects an increment of attentional control in HP-EH-EVOO group which was reflected to improved neuropsychological estimates. Moreover, we reported a reduction of theta signal spectrum in both conditions mostly for the HP-EH-EVOO and secondly for the MP-EVOO underlined by large and medium effect size, correspondingly. Group-averaged ΔNI^θ^ was decreased after the intervention in both conditions and across groups.

Global alpha synchrony is an index of a healthy resting wakefulness where a subject can process any information receiving from the real world (Klimesch,1999). Alpha desynchronization can be realized a) via the activation of the visual system (eyes open compared to eyes closed condition) mediated by the reticular system (Volavka et al.,1967) and also b) due to changes of the cortical network and thalamo-cortical communication (Schurmann and Basar, 2001; Fisch, 2006). We observed a reduction of alpha signal spectrum in both conditions mostly for the HP-EH-EVOO and secondly for the MP-EVOO underlined by large and medium effect size, correspondingly. Group-averaged ΔNI^α^ were higher for eyes-open condition compared to eyes-closed in the three groups. ΔNI^α^ was decreased after the intervention in both conditions and mostly for HP-EH-EVOO group who showed significantly higher post-intervention reduction compared to the rest two groups. A recent study reported that EEG alpha power was significantly enhanced during mind wandering compared to a cognitive task underlying the its importance to explore temporal fluctuations of mind wandering (Compton et al., 2019).

Beta frequency power is involved at EEG resting-state networks in a healthy population (Mantini et al., 2007). Beta frequency has been linked to lower MMSE scores (Stam et al., 2003) while is part of an integrative biomarker that can predict the progression to AD (Poil et al., 2013). Complementary, a reduced beta power has been observed during mind wandering compared to cognitive tasks in a healthy group (Van Son et al., 2019). Here,we observed a reduction of beta signal spectrum in both conditions mostly for the HP-EH-EVOO and secondly for the MP-EVOO underlined by large and medium effect size, correspondingly. The reduction of ΔNI^β^ was higher for HP-EH-EVOO group compared to MeDi group in eyes-closed condition.

Fundamental changes of signa power and connectivity were observed in al the studied brain disorders/diseases with the direction of changes to be associated with the task (Basar et al., 2013). For that reason, brain oscillations can be used as possible biomarkers in clinical populations studies and also for the evaluation of treatment and intervention protocols. It is highly important to compare similar conditions and especially for resting-state conditions, eyes-closed should be trated as an arousal baseline while eyes-open condition as an activation baseline.

The fluctuation of dominant coupling modes quantified with FI and the related pre-post difference estimated with ***ΔFI***^***DoCM***^ showed an improvement in the three groups and in both conditions. We observed higher ***ΔFI***^***DoCM***^ values for HP-EH-EVOO and MP-EVOO in eyes-open compared to eyes-closed condition even though the findings didn’t reach significant level.

HP-EH-EVOO group showed significant higher post-intervention reduction of the NI level in total and in α in both conditions. Interestingly, group-averaged ΔNI^α^ were higher for eyes-open condition compared to eyes-closed in the three groups. Complementary, we observed important to mention patterns for the rest of potential modulating frequencies. ΔNI^δ^ was increased after the intervention in both conditions and across groups while ΔNI^θ,β^ were both reduced after the intervention in both conditions and across groups. We detected the following significant differences between groups : i) the increment of ΔNI^δ^ was higher for MeDi compared to HP-EH-EVOO group in both conditionsand ii) the reduction of ΔNI^β^ was higher for HP-EH-EVOO group compared to MeDi group in eyes-closed condition.

Following a multi-linear regression analysis with ΔNI^Total or α^ as responses and the Δ differences of eight neuropsychological assessments as potential predictors untangled a link between reduction of ΔNI^Total^ with Δ differences of eight neuropsychological variables. This is the very first study that reported a link between aberrant changes of DoCM quantified via the NI with neuropsychological estimates due to an intervention protocol.

The combination of the main key findings which are a) the increment of *ΔFI*^*DoCM*^ and the reduction of post-intervention NI level in total and in α showed with negative values of ΔNI^total,α^ can support the following statement: Intervention protocol reduced the excitation of information flow expressed with the ratio of CFC over WFC which is driven mostly by α modulating frequency. A high *ΔFI*^*DoCM*^ with a less involvement of CFC (reduction of NI) mostly modulating by α frequency means that dominant coupling modes fluctuate faster and are represented in a higher proportion over WFC compared to CFC.

This study complements the positive outcome of the first study that used HP-EH-EVOO. We further validated the improved cognitive measures published in the original MICOIL study (Tsolaki et al., 2020) by analysing the EEG resting-state recordings of a specific subgroup of subjects. Signal spectrum analysis and dynamic functional connectivity analysis was followed. Adopting our DoCM model, we explored how an intervention protocol tailored to MCI subjects can alter the multiplexity of dynamic functional connectivity in a positive direction (Buzsáki and Watson, 2012). The whole approach proved more sensitive to detect alterations of brain activity and connectivity compared to the trivial neuropsychological testing even on our small sample. The combination of neuropsychological estimates and EEG recordings is a promising low-cost and non-invasive monitoring assessment of inviduals at risk (Babiloni et al., 2010; Lizio et al., 2011)

Our study is novel and unique in both the intervention protocol and the adopted analytic pathway. However, we can report two basic limitations. The first one refers to our analysis using scalp EEG sensors. For that reason and in order to avoid and misleading interpretations of our findings in a local level, we reported our estimates across the whole EEG space. It would be very interesting to follow a similar protocol employing a high-density EEG system and work on virtual cortical space instead of scalp surface level. The second limitation is linked to the acceptable but small number of participants. Power analysis supported our findings but we could report that a higher number of participants will definitely be on the right direction.

## 5. Conclussions

In the present study, we evaluated for the very first time the effect of Greek High Phenolic Early Harvest Extra Virgin Olive Oil (HP-EH-EVOO) versus Moderate Phenolic (MP-EVOO) and Mediterranean Diet (MeDi) in people with MCI via the EEG resting-state analysis. Our analysis was unique in terms of combining both intra and cross-frequency interactions simultaneously under the DoCM model. We reported for the very first time a reconfiguration of phase driven DoCM in MCI subjects that followed a dietary protocol while this reconfiguration was more prominent for the HP-EH-EVOO group. Signal power reduced across the spectrum with most important findings up to 15 Hz for HP-EH-EVOO and MP-EVOO groups. Our analytic pathway can assist researchers of how they have to evaluate their intervention protocols tailored to MCI subjects but also in other target groups. Further analysis is needed to link our DoCM findings with the intervention protocol in a larger sample.

## Data Availability

The dataset is not available to the public.

## Author’s Contribution

**SID:** Conceptualization; Data curation; Formal analysis; Methodology; Writing - original draft

**CL** : EEG Data curation; Formal analysis; EEG Data collection. A.Ch,T.

EL. All Data collection (Demographics, Genetics, MRI, Blood examination

MC. Neuropsychological Examination

MT. Organization the methodology of the study, neurological examination of the patients, decision for the inclusion-exclusion criteria and final editing of the study

## Acknowledgement

This work has been supported by the Greek Association of Alzheimer’s Disease and Related Disorders (GAADRD), theWorld Olive Centre for Health (WOCH) and Yanni’s Olive Grove company for the donation of HP-EH-EVOO: Yanni’s Fresh and MPEVOO: Yanni’s Selected. In addition, we would like to thank Panagiotis Diamantakos and Aimilia Riga kou for technical support in the analysis of EVOO, Hadar Halivni for editing the English language, and all clinicians who contributed to this study, as well as patients who took part in it. SD was supported by a MRC grant MR/K004360/1 (Behavioural and Neurophysiological Effects of Schizophrenia Risk Genes: A Multi-locus, Pathway Based Approach) and a MARIE-CURIE COFUND EU-UK Research Fellowship.

## Footnotes

1 https://clinicaltrials.gov/ct2/show/NCT03362996

## References

Abuznait AH, Qosa H, Busnena BA, El Sayed KA, Kaddoumi A. Olive-oil-derived oleocanthal enhances β-amyloid clearance as a potential neuroprotective mechanism against Alzheimer’s disease: in vitro and in vivo studies. ACS Chem Neurosci. 2013 Jun 19;4(6):973–82

Andrich K, Bieschke J. The Effect of (-)-Epigallo-catechin-(3)-gallate on Amyloidogenic Proteins Suggests a Common Mechanism. Adv Exp Med Biol. 2015;863:139–61. doi: 10.1007/978-3-319-18365-7_7.

Antonakakis M, Dimitriadis SI, Zervakis M, Micheloyannis S, Rezaie R, Babajani-Feremi A, Zouridakis G, Papanicolaou AC. Altered cross-frequency coupling in resting-state MEG after mild traumatic brain injury. Int J Psychophysiol. 2016 Apr;102:1–11.

Antonakakis M, Dimitriadis SI, Zervakis M, Papanicolaou AC, Zouridakis G. Reconfiguration of dominant coupling modes in mild traumatic brain injury mediated by δ-band activity: A resting state MEG study. Neuroscience. 2017 Jul 25;356:275–286.

Ayissi VB, Ebrahimi A, Schluesenner H. Epigenetic effects of natural polyphenols: a focus on SIRT1-mediated mechanisms. Mol Nutr Food Res. 2014 Jan;58(1):22–32.

Babiloni C, Visser PJ, Frisoni G, De Deyn PP, Bresciani L, Jelic V, Nagels G, Rodriguez G, Rossini PM, Vecchio F, Colombo D, Verhey F, Wahlund LO, Nobili F. Cortical sources of resting EEG rhythms in mild cognitive impairment and subjective memory complaint. Neurobiol Aging. 2010 Oct;31(10):1787–98.

Barry RJ, Clarke AR, Johnstone SJ, Magee CA, Rushby JA. EEG differences between eyes-closed and eyes-open resting conditions. Clin Neurophysiol. 2007 Dec;118(12):2765–73.

Başar E, Başar-Eroğlu C, Güntekin B, Yener GG. Brain’s alpha, beta, gamma, delta, and theta oscillations in neuropsychiatric diseases: proposal for biomarker strategies. Suppl Clin Neurophysiol. 2013;62:19–54.

Brambati SM, Belleville S, Kergoat MJ, Chayer C, Gauthier S, Joubert S. Single- and multiple-domain amnestic mild cognitive impairment: two sides of the same coin? Dement Geriatr Cogn Disord. 2009;28(6):541–49.

Buzsáki G, Watson BO. Brain rhythms and neural syntax: implications for efficient coding of cognitive content and neuropsychiatric disease. Dialogues Clin Neurosci. 2012 Dec;14(4):345–67.

Chen CC, Henson RN, Stephan KE, Kilner JM, Friston KJ. Forward and backward connections in the brain: a DCM study of functional asymmetries. Neuroimage 2009;45:453–62.

Chun-Chuan Chen, James M. Kilner, Karl J. Friston, Stefan J. Kiebel, Rohit K. Jolly, Nick S. Ward. Nonlinear Coupling in the Human Motor System. Journal of Neuroscience 23 June 2010;30(25):8393–8399

Cohen G. Oxy-radical toxicity in catecholamine neurons. Neurotoxicology. 1984 Spring;5(1):77–82.

Compton, R.J., Gearinger, D. & Wild, H. The wandering mind oscillates: EEG alpha power is enhanced during moments of mind-wandering. Cogn Affect Behav Neurosci 2019;19:1184–1191 (2019).

Declerck K, Szarc vel Szic K, Palagani A, Heyninck K, Haegeman G, Morand C, Milenkovic D, Vanden Berghe W. Epigenetic control of cardiovascular health by nutritional polyphenols involves multiple chromatin-modifying writer-reader-eraser proteins. Curr Top Med Chem. 2016;16(7):788–806.

Dimitriadis SI, Salis C, Tarnanas I, Linden DE. Topological Filtering of Dynamic Functional Brain Networks Unfolds Informative Chronnectomics: A Novel Data-Driven Thresholding Scheme Based on Orthogonal Minimal Spanning Trees (OMSTs). Front Neuroinform. 2017 Apr 26;11:28.

Dimitriadis SI, Antonakakis M, Simos P, Fletcher JM, Papanicolaou AC. Data-Driven Topological Filtering Based on Orthogonal Minimal Spanning Trees: Application to Multigroup Magnetoencephalography Resting-State Connectivity. Brain Connect. 2017 Dec;7(10):661–670.

Dimitriadis SI, Salis CI. Mining Time-Resolved Functional Brain Graphs to an EEG-Based Chronnectomic Brain Aged Index (CBAI). Front Hum Neurosci. 2017 Sep 7;11:423.

Dimitriadis SI, López ME, Bruña R, Cuesta P, Marcos A, Maestú F, Pereda E. How to Build a Functional Connectomic Biomarker for Mild Cognitive Impairment From Source Reconstructed MEG Resting-State Activity: The Combination of ROI Representation and Connectivity Estimator Matters. Front Neurosci. 2018;1:12:306.

Dimitriadis SI, Routley B, Linden DE, Singh KD. Reliability of Static and Dynamic Network Metrics in the Resting-State: A MEG-Beamformed Connectivity Analysis. Front Neurosci. 2018;12:506.

Dimitriadis SI, Simos PG, Fletcher JΜ, Papanicolaou AC. Aberrant resting-state functional brain networks in dyslexia: Symbolic mutual information analysis of neuromagnetic signals. Int J Psychophysiol. 2018 Apr;126:20–29.

Dimitriadis SI. Complexity of brain activity and connectivity in functional neuroimaging. J Neurosci Res. 2018 Nov;96(11):1741–1757.

Dimitriadis SI, Simos PG, Fletcher JΜ, Papanicolaou AC. Typical and Aberrant Functional Brain Flexibility: Lifespan Development and Aberrant Organization in Traumatic Brain Injury and Dyslexia. Brain Sci. 2019 Dec 16;9(12):380.

Dimitriadis SI (2020). Reconfiguration of αmplitude driven dominant coupling modes (DoCM) mediated by α-band in adolescents with schizophrenia spectrum disorders. Progress in Neuro-Psychopharmacology and Biological Psychiatry In Press. https://doi.org/10.1016/j.pnpbp.2020.110073

Eastwood MA. Interaction of dietary antioxidants in vivo: How fruit and vegetables prevent disease? QJM Mon. J. Assoc. Phys. 1999;92:527–530.

Féart, C.; Samieri, C.; Barberger-Gateau, P. Mediterranean diet and cognitive function in older adults. Curr. Opin. Clin. Nutr. Metab. Care 2010, 13, 14–18.

Féart C, Samieri C, Allès B, Barberger-Gateau P. Potential benefits of adherence to the Mediterranean diet on cognitive health. Proc. Nutr. Soc. 2013;72:140–152.

Finkel T, Holbrook NJ. Oxidants, oxidative stress and the biology of ageing. Nature. 2000;408:239–247.

Fisch NJ. Alpha channeling in mirror machines. Phys. Rev. Lett. 2006;97:225001.

Goszcz K, Duthie GG, Stewart D, Leslie SJ, Megson IL. Bioactive polyphenols and cardiovascular disease: Chemical antagonists, pharmacological agents or xenobiotics that drive an adaptive response? Br. J.Pharmacol. 2017;174:1209–1225.

Grossi C, Rigacci S, Ambrosini S, Dami TE, Luccarini I, Traini C, Failli P, Berti A, Casamenti F, Stefani M, et al. The polyphenol oleuropein aglycone protects TgCRND8 mice against A_ plaque pathology. PLoS ONE 2013;8:e71702.

Harmony T. The functional significance of delta oscillations in cognitive processing. Front Integr Neurosci. 2013;7:83.

Herrera CG, Cadavieco MC, Jego S, Ponomarenko A, Korotkova T, Adamantidis A. Hypothalamic feedforward inhibition of thalamocortical network controls arousal and consciousness. Nat Neurosci. 2016 Feb;19(2):290–8.

Hollman PC, Katan MB. Health effects and bioavailability of dietary flavonols. Free Radic. Res. 1993;31S75–S80.

Hyafil A, AL Giraud, L Fontolan, B Gutkin. Neural cross-frequency coupling: connecting architectures, mechanisms, and functions Trends in Neurosciences 2015;38:725–740.

Karakaya T, Fußer F, Schröder J, Pantel J. Pharmacological Treatment of Mild Cognitive Impairment as a Prodromal Syndrome of Alzheimer’s Disease. Curr Neuropharmacol. 2013;11(1):102–108.

Kim HS, Quon MJ, Kim JA. New insights into the mechanisms of polyphenols beyond antioxidant properties; lessons from the green tea polyphenol, epigallocatechin 3-gallate. Redox Biol. 2014;2:187–195.

Klimesch W. EEG alpha and theta oscillations reflect cognitive and memory performance: a review and analysis. Brain. Res. Brain. Res. Rev. 1999;29:169–195.

Knyazev G. EEG delta oscillations as a correlate of basic homeostatic and motivational processes. Neurosci. Biobehav. Rev. 2012;36:677–695

Lizio R, Vecchio F, Frisoni GB, Ferri R, Rodriguez G, Babiloni C. Electroencephalographic rhythms in Alzheimer’s disease. Int J Alzheimers Dis. 2011;2011:927573.

López-Miranda J, Pérez-Jiménez F, Ros E, De Caterina R, Badimón L, Cova MI, Escrich E, Ordovás JM, Soriguer F, Abiá R et al. Olive oil and health: Summary of the II international conference on olive oil and health consensus report, Jaén and Córdoba (Spain) 2008. Nutr. Metab. Cardiovasc. Dis. 2010;20:284–294.

Lyras L,Cairns NJ, Jenner A, Jenner P, Halliwell B. An assessment of oxidative damage to proteins, lipids, and DNA in brain from patients with Alzheimer’s disease. J Neurochem. 1997; 68: 2061–2069

Mantini, Perrucci MG, Del Gratta C, Romani GL, Corbetta M. Electrophysiological signatures of resting state networks in the human brain. Proc. Natl. Acad. Sci. U.S.A. 2007;104:13170–13175.

Marimpis D, Dimitriadis SI and Goebel R. A Multiplex Connectivity Map of Valence-Arousal Emotional Model. IEEE Access, 2020;8:170928–170938

Martínez-Lapiscina EH, Clavero P, Toledo E, San Julián B, Sanchez-Tainta A, Corella D, Lamuela-Raventós RM, Martínez JA, Martínez-Gonzalez MA. Virgin olive oil supplementation and long-term cognition: The PRED IMED-NAVARRA randomized, trial. J Nutr Health Aging 2013;17:544–552.

McCord JM. The evolution of free radicals and oxidative stress. Am J Med. 2000;1:108(8):652–9.

Mesulam MM, Mufson EJ. Insula of the old world monkey III: efferent cortical output and comments on function. J. Comp. Neurol. 1982;212:38–52

Middleton E Jr. Effect of plant flavonoids on immune and inflammatory cell function. Adv. Exp. Med. Biol. 1998;439:175–182.

Nunez PL and Srinivasan R (2006) Electric Fields of the Brain: The Neurophysics of EEG, 2nd Edition, New York: Oxford University Press.

Otaegui-Arrazola A, Amiano P, Elbusto A, Urdaneta E, Martínez-Lage P. Diet, cognition, and Alzheimer’s disease: food for thought. Eur J Nutr. 2014 Feb;53(1):1–23.

Petersen RC, Smith GE, Waring SC, Ivnik RJ, Tangalos EG, Kokmen E. Mild cognitive impairment: clinical characterization and outcome. Arch. Neurol. 1999;56:303–308.

Pitozzi V, Jacomelli M, Catelan D, Servili M, Taticchi A, Biggeri A, Dolara P, Giovannelli L. Long-term dietary extra-virgin olive oil rich in polyphenols reverses age-related dysfunctions in motor coordination and contextual memory in mice: Role of oxidative stress. Rejuvenation Res. 2012;15:601–612.

Pitt J, Roth W, Lacor P, et al. Alzheimer’s-associated Abeta oligomers show altered structure, immunoreactivity and synaptotoxicity with low doses of oleocanthal. Toxicol Appl Pharmacol. 2009;240(2):189–197.

Poil S S, de Haan W, van der Flier WM, Mansvelder HD, Scheltens P, Linkenkaer-Hansen K. Integrative EEG biomarkers predict progression to Alzheimer’s disease at the MCI stage. Front. Aging Neurosci. 2013;5:58.

Qosa H, Mohamed LA, Batarseh YS, Alqahtani S, Ibrahim B, LeVine H, Keller JN, Kaddoumi A. Extra-virgin olive oil attenuates amyloid-_ and tau pathologies in the brains of TgSwDI mice. J Nutr Biochem 2015;26:1479–1490.

Rigacci S, Guidotti V, Bucciantini M, Nichino D, Relini A, Berti A, Stefani MA. (1–42) aggregates into non-toxic amyloid assemblies in the presence of the natural polyphenol oleuropein aglycon. Curr. Alzheimer Res. 2011;8:841–852.

Sauseng PB, Griesmayr R, Freunberger W. KlimeschControl mechanisms in working memory: a possible function of EEG theta oscillations.Neurosci. Biobehav. Rev. 2010;34:1015–1022

Scarmeas N, Stern Y, Tang MX, Mayeux R, Luchsinger JA. Mediterranean diet and risk for Alzheimer’s disease. Ann. Neurol. 2006;59:912–921.

Scarmeas N, Stern Y, Mayeux R, Manly JJ, Schupf N, Luchsinger JA. Mediterranean diet and mild cognitive impairment. Arch Neurol 2009;66:216–225.

Scarmeas N, Luchsinger JA, Stern Y, Gu Y, He J, De Carli C, Brown T, Brickman AM Mediterranean diet and magnetic resonance imaging-assessed cerebrovascular disease. Ann. Neurol. 2011;69:257–268.

Scarmeas N, Anastasiou CA, Yannakoulia M. Nutrition and prevention of cognitive impairment. Lancet Neurol. 2018;17(11):1006–1015.

Servili M, Montedoro G. Contribution of phenolic compounds to virgin olive oil quality. Eur. J. Lipid Sci. Technol. 2002;104:602–613.

Servili M, Sordini B, Esposto S, Urbani S, Veneziani G, Di Maio I, Selvaggini R, Taticchi A. Biological activities of phenolic compounds of extra virgin olive oil. Antioxidants 2013;3:1–23.

Schurmann M, Basar E. Functional aspects of alpha oscillations in the EEG. Int. J. Psychophysiol. 2001;39;151–158.

Singh M, Arseneault M, Sanderson T, Murthy V, Ramassamy C. Challenges for research on polyphenols from foods in Alzheimer’s disease: Bioavailability, metabolism, and cellular and molecular mechanisms. J. Agric. Food Chem. 2008;56:4855–4873.

Sofi F, Abbate R, Gensini GF, Casini A. Accruing evidence on benefits of adherence to the Mediterranean diet on health: An updated systematic review and meta-analysis. Am J Clin Nutr 2010; 92:1189–1196.

Stam CJ, van der Made Y, Pijnenburg YA, Scheltens P. EEG Synchronization in Mild Cognitive Impairment and Alzheimer’s Disease. Acta Neurol. Scand. 2003;108: 90–96.

Tsolaki M, Karathanasi E, Lazarou I, Dovas K, Verykouki818 E, Karacostas A, Georgiadis K, Tsolaki A, Adam K, Kompatsiaris I, Sinakos Z. Efficacy and safety of Crocus sativus L. in patients with mild cognitive impairment: One year single-blind randomized, with parallel groups, clinical trial. J Alzheimers Dis 2016;54:129–133.

Tsolaki M, Lazarou E, Kozori M, Petridou N, Tabakis I, Lazarou I, Karakota M, Saoulidis I, Melliou E, Magiatis P. A Randomized Clinical Trial of Greek High Phenolic Early Harvest Extra Virgin Olive Oil in Mild Cognitive Impairment: The MICOIL Pilot Study. J Alzheimers Dis. 2020;78(2):801–817.

Vlachos GS, Scarmeas N. Dietary interventions in mild cognitive impairment and dementia. Dialogues Clin Neurosci. 2019;21(1):69–82.

Volavka J, Matousek M, Roubicek J. Mental arithmetic and eye opening. An EEG frequency analysis and GSR study. Electroencephalogr. Clin. Neurophysiol. 1967;22:174–176.

van Son D, De Blasio FM, Fogarty JS, Angelidis A, Barry RJ, Putman P. Frontal EEG theta/beta ratio during mind wandering episodes. Biological Psychology, 2019;140:19–27.

von Bernhardi R, Eugenin J. Alzheimer’s disease: Redox dysregulation as a common denominator for diverse 851 pathogenicmechanisms. AntioxidRedoxSignal 2012;16, 974–1031

Weyand TG, Boudreaux M, Guido W. Burst and tonic response modes in thalamic neurons during sleep and wakefulness. J Neurophysiol. 2001 Mar;85(3):1107–18.

Winblad B, Palmer K, Kivipelto M, Jelic V, Fratiglioni L, Wahlund LO, Nordberg A, Bäckman L, Albert M, Almkvist O, Arai H, Basun H, Blennow K, de Leon M, DeCarli C, Erkinjuntti T, Giacobini E, Graff C, Hardy J, Jack C, Jorm A, Ritchie K, van Duijn C, Visser P, Petersen RC. Mild cognitive impairment-beyond controversies, towards a consensus: report of the International Working Group on Mild Cognitive Impairment. J. Intern. Med. 2004;256:240–246.

